# Development of an LLM Pipeline Surpassing Physicians in Cardiovascular Risk Score Calculation

**DOI:** 10.1101/2025.11.11.25340002

**Authors:** Tobias Roeschl, Marie Hoffmann, Axel Unbehaun, Henryk Dreger, Gerhard Hindricks, Volkmar Falk, Ran Balicer, Radu Tanacli, Felix Hohendanner, Alexander Meyer

**Affiliations:** Deutsches Herzzentrum der Charité, Department of Cardiology, Angiology and Intensive Care Medicine, Charitéplatz 1, 10117 Berlin, Germany; Charité – Universitätsmedizin Berlin, corporate member of Freie Universität Berlin and Humboldt Universität zu Berlin, Charitéplatz 1, 10117 Berlin, Germany; Charité – Universitätsmedizin Berlin, Institute for Artificial Intelligence in Medicine, Invalidenstraße 120, 10115 Berlin, Germany; Berlin Institute of Health at Charité – Universitätsmedizin Berlin, Charitéplatz 1, 10117 Berlin, Germany; DZHK (German Centre for Cardiovascular Research), partner site Berlin, Berlin, Germany; Deutsches Herzzentrum der Charité, Department of Cardiothoracic and Vascular Surgery, Augustenburger Platz 1, 13353 Berlin, Germany; Deutsches Herzzentrum der Charité, Department of Cardiology, Angiology and Intensive Care Medicine, Augustenburger Platz 1, 13353 Berlin, Germany; Translational Cardiovascular Technologies, Institute of Translational Medicine, Department of Health Sciences and Technology, Swiss Federal Institute of Technology, 8092 Zürich, Switzerland; Innovation Division, Clalit Health Services, Clalit Research Institute, Tel Aviv, Israel; Berlin Institute for the Foundations of Learning and Data – TU Berlin, Berlin, Germany

## Abstract

**Background:** Risk scores are essential to evidence-based cardiovascular care, but manual calculation is labor-intensive and error-prone. Large language models (LLMs) could automate this process, yet LLMs are limited by their propensity for calculation errors and factual hallucinations. Pipelines that separate LLM-based data extraction from deterministic score computation may improve reliability and transparency.

**Methods:** We conducted a retrospective diagnostic study at a quaternary heart center in Germany (January 2020 – July 2023). Patients with atrial fibrillation (n=179) from an ablation registry and patients with severe aortic stenosis (n=76) evaluated by a heart team were included. Five LLMs (DeepSeek-R1, Qwen3, GPT-4 Turbo, Llama 3.1, and PaLM 2) were tested in standalone and pipeline configurations to compute HAS-BLED, CHA₂DS₂-VASc, and EuroSCORE II scores from routine clinical reports. Accuracy was assessed by comparing predictions to expert-adjudicated ground truth, using root mean squared error (RMSE), Krippendorff’s alpha for categorical agreement, and calibration analysis.

**Results:** Pipeline-generated scores showed substantially higher agreement with expert adjudication than standalone LLMs and treating clinicians (mean Krippendorff’s alpha: 0.79 vs 0.32 vs 0.31) and demonstrated superior calibration. The Qwen3-based pipeline, achieved the highest accuracy with lower RMSEs than clinicians for HAS-BLED (0.20 vs 0.87), CHA₂DS₂-VASc (0.53 vs 1.08), and EuroSCORE II (1.99 vs 2.05).

**Conclusion:** LLM-based pipelines enable accurate, well-calibrated, and scalable cardiovascular risk score computation from unstructured real-world clinical data, outperforming clinicians and standalone LLMs with the potential to reduce clinician workload and support evidence-based care.

## Introduction

Clinical risk scores are foundational instruments in medicine, translating complex patient data into actionable risk estimates that guide therapeutic decisions and support guideline-adherent care. As medicine becomes increasingly data-driven^1^, accurate calculation of these scores is vital. Typically, physicians must manually extract variables from unstructured electronic health records – a workflow that is both laborious and susceptible to error.

Large language models (LLMs) could automate such text-heavy clinical tasks, yet their application to quantitative risk scoring is fundamentally undermined by well-documented failure modes. These models are prone to factual and contextual hallucinations^2^ – generating confident but incorrect outputs – and often exhibit poor arithmetic reasoning^3–5^, limitations that are particularly hazardous where patient safety and guideline adherence are at stake. While recent work^6^ has shown that integrating LLMs with external calculators can improve numerical accuracy, a critical translational gap persists. Previous studies have relied on synthetic vignettes or concise case reports, which do not reflect the complexity and noise of authentic clinical documents such as multi-page discharge letters or imaging reports^3,6–8^. Performance of these systems in a real-world clinical setting therefore remains unknown.

To date, no study has validated an LLM-based system for risk score calculation on authentic, unstructured patient reports, or rigorously benchmarked against physicians.

In our study, we aimed to address these gaps by evaluating how accurately standalone LLMs and LLM-based pipelines compute cardiovascular risk scores from routine clinical text reports compared to treating physicians. The LLM-based pipeline we developed integrates the language-understanding capabilities of LLMs with the reliability of deterministic computation. It comprises three stages: (1) expert-curated, guideline-concordant score definitions, (2) LLM-driven extraction of relevant data from original clinical reports, and (3) a separate, formula-based engine for the final score computation.

We conducted this comparison using three widely adopted cardiovascular risk scores – CHA₂DS₂-VASc⁹, HAS-BLED¹⁰, and EuroSCORE II¹¹– across two patient cohorts. Our findings show that pipeline models achieve superior accuracy and calibration, outperforming both standalone LLMs and treating physicians. This demonstrates its potential as a robust and scalable framework for automated clinical risk assessment.

## Methods

### Study Design and Patient Cohorts

Our retrospective study included two distinct patient cohorts from a single quaternary care heart center. The first cohort included 179 of 182 consecutively screened adult patients from an in-house registry who underwent catheter ablation for atrial fibrillation (AF) between January 2020 and July 2023, each with a digitized discharge letter. The second cohort included 76 of 90 adults with severe aortic stenosis (AS) evaluated by one of our institutional heart teams during 2022, each with comprehensive digitized documentation. For the AS cohort, sufficient documentation was defined as the availability of at least one physician letter, echocardiography report, cardiac catheterization report, computed tomography (CT) report, and a heart team protocol. Patient characteristics for both cohorts are detailed in Supplementary Table 1 and Supplementary Table 2. The study protocol was approved by the research ethics committee of Charité – Universitätsmedizin Berlin (EA2/183/23).

### Data Acquisition and Preparation

All clinical documents were extracted as Portable Document Format (PDF) files from the hospital information system and converted into plain text using optical character recognition. For each patient in the AF cohort, the physician letter following the ablation procedure was used as the input document. For the AS cohort, a comprehensive single document was created for each patient by concatenating their two most recent discharge letters, the invasive coronary angiography report, echocardiography report, and CT scan report. This was done to simulate a complete chart review.

All reports were manually anonymized by replacing identifiable information with synthetic data, while preserving the original structure and clinical complexity. To maintain temporal relationships between events, all dates for a given patient were shifted by a randomly sampled offset.

We manually extracted risk scores documented by treating clinicians in discharge letters and heart team protocols to establish a reference standard for benchmarking. These values were then removed from the documents provided to the LLMs to prevent data leakage.

### Model Architectures and Prompting Strategies

Figure 1 provides a comprehensive overview of the two approaches evaluated. In the standalone LLM-approach, we instructed LLMs to calculate scores directly from clinical reports with reference only to the original score publications. In a single, comprehensive prompt, the full clinical document was provided alongside the instruction to calculate the complete risk score according to the respective original publication and output both the final score and all contributing score components in a structured JSON format.

**Figure 1:**
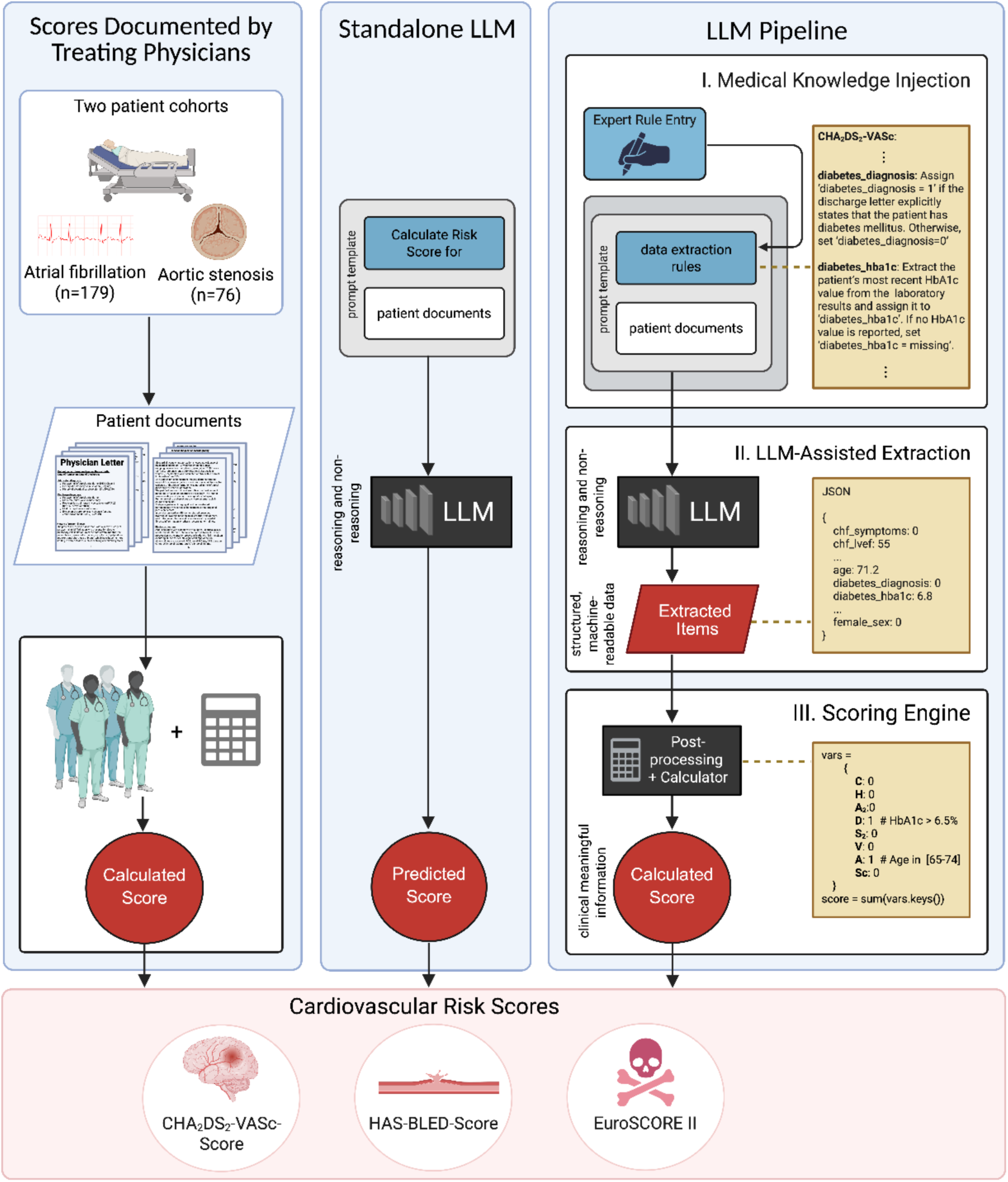
Experimental design. Three experimental arms evaluate risk score derivation in two patient cohorts based on original patient documents. Left: Risk scores treating physicians had manually calculated during clinical practice. Center: In the standalone LLM approach, a single prompt is used to directly predict the risk score end-to-end from patient documents. Right: In LLM pipeline, a three-stage process is applied: Stage I, Medical knowledge is incorporated into prompts for granular data extraction; Stage II, the LLM outputs structured values for each variable; and Stage III, a deterministic scoring engine derives aggregated component values and applies validated formulas to compute the final risk score.

In the pipeline approach, risk score computation was organized into three stages: (1) provision of score component definitions, including surrogate parameters, enhanced by domain-specific knowledge incorporated into all prompt templates to integrate clinical expertise (e.g., symptom interpretation criteria) and specifications from the original score publications; (2) LLM-based extraction of structured data from clinical reports; and (3) deterministic calculation of the final score. The inclusion of surrogate parameters beyond the classical score components enabled a more comprehensive assessment of patient risk. For example, the models were prompted to use the most recent HbA1c value to infer the presence of diabetes mellitus and assign a corresponding point in the CHA₂DS₂-VASc score, even when the diagnosis was not explicitly mentioned in the discharge letter. To improve extraction accuracy, we issued a separate query for each individual score component. A core prompt template was used for each score component across all models. Prompt templates are provided in the Supplementary material, and representative examples are shown in Figure 2. As in the standalone LLM approach, we instructed the LLMs to output structured output in JSON format. We developed a deterministic scoring engine, coded in Python, that used a rule-based approach to further process the extracted data and apply logical and arithmetic operations to compute the final score.

**Figure 2:**
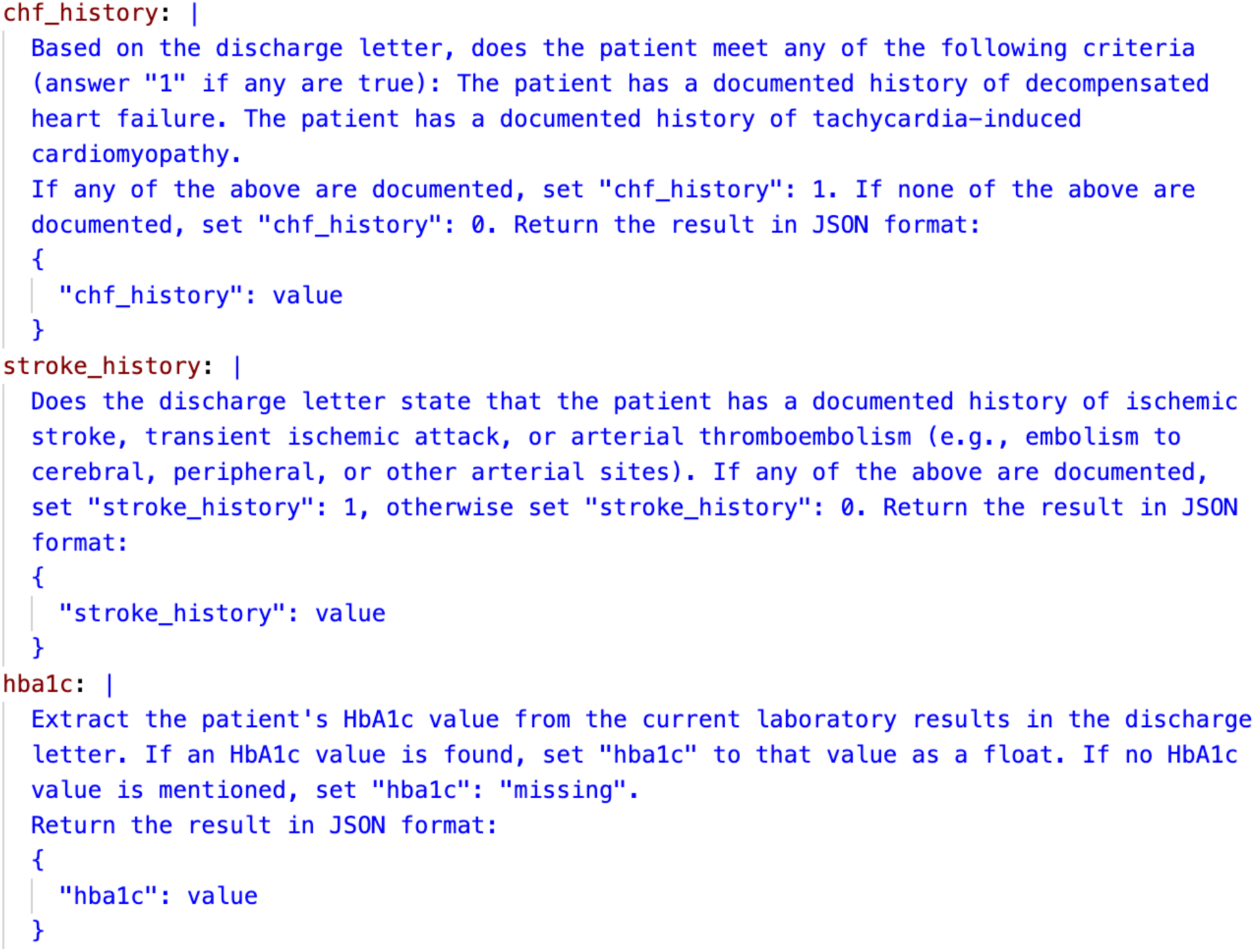
Prompt templates. Representative examples of the prompt templates used in the CHA_2_DS_2_-VASc scoring pipeline.

In both approaches we tested five different LLMs - two optimized for reasoning (DeepSeek-R1, Qwen3-235B-A22B) and three leading models (GPT-4 Turbo, Llama 3.1, PaLM 2). All prompts were executed in a zero-shot configuration with the temperature parameter set to zero to minimize output variation and enhance reproducibility.

### Ground Truth Adjudication Process

To establish a reference standard for all risk scores, two senior resident cardiologists independently reviewed all patient documents for both cohorts, blinded to the scores documented by the treating physicians. The cardiologists used a standardized data collection form detailing the definitions for every component of the HAS-BLED, CHA_2_DS_2_-VASc, and EuroSCORE II scores, as specified in their original publications (Supplementary Table 3). Raters were instructed to use the most recent value for any duplicate entries and to assume normal reference ranges for missing laboratory values. A standardized mapping was used for qualitative descriptions of left-ventricular ejection fraction (e.g., ‘moderately impaired’ was mapped to 35%). Any disagreements between the two primary raters were resolved by a third, independent senior cardiologist to reach a final consensus for each score component. Ground truth patient ages were programmatically calculated based on the date of birth and either the date of discharge or the date of the heart team meeting. Ground truth scores were calculated from these components using the score points and regression coefficients reported in the original publications, respectively.

To evaluate agreement between clinical raters during the establishment of ground truth values for score components, we employed Krippendorff’s alpha, using the nominal or ordinal variant as appropriate based on the measurement level of each variable. Due to the instability or inapplicability of these metrics for instances with sparse data, we additionally reported raw agreement as the proportion of identical ratings between raters. Once consensus ground truth scores were established, we evaluated the performance of standalone LLMs, pipeline models and physician-derived scores against these reference standards.

### Performance metrics and Statistical Analysis

Patient characteristics were reported as counts and percentages for binary variables, as means with 95% confidence intervals for normally distributed continuous variables, and as medians with interquartile ranges for non-normally distributed variables. Normality was assessed using the Shapiro–Wilk test.

We evaluated agreement between risk scores derived by the standalone LLMs, the pipeline models, and treating physicians, and the ground truth scores across clinically relevant score categories. Specifically, we used the following threshold groupings: <2 vs ≥2 for CHA_2_DS_2_-VASc^12^, <3 vs ≥3 for HAS-BLED^10^, and <4%, 4–8%, >8% for EuroSCORE II^13^. Agreement across these categories was assessed using Krippendorff’s alpha, excluding instances with missing data. Prediction accuracy was quantified using root mean squared error (RMSE) between each derived score and the ground truth.

Calibration of physician-derived scores and pipeline model-derived scores was evaluated using kernel density estimation (KDE) plots, with ground truth scores on the x-axis and predicted scores on the y-axis. The diagonal dashed line indicates perfect calibration, with density contours closer to this line reflecting better calibration.

Per-component accuracies, defined as the proportion of correctly assigned values for each score component, were visualized using radar plots. For the age variable, predictions within ±1 year were counted as correct for this metric.

Frequency distributions of LLM-derived scores were visualized using histograms to detect output patterns suggestive of repetitive responses. All statistical analyses were conducted using Python version 3.9, scikit-learn 1.6, numpy 1.26, and krippendorff 0.8.

## Results

### Standalone LLMs do not ensure valid risk assessments for complex risk scores

Applying these evaluation metrics, the standalone LLMs tasked with end-to-end score calculation produced inconsistent and often erroneous results. While some LLMs matched the accuracy of treating physicians for the simpler additive HAS-BLED and CHA_2_DS_2_-VASc scores, all models failed to correctly calculate the more complex EuroSCORE II, which is derived from a logistic regression model applying the logistic function to a weighted linear combination of clinical variables. All LLMs tested markedly overestimated this score. This failure was evident in the extremely high RMSE values and poor agreement for EuroSCORE II scores derived by standalone LLMs (Figure 3 and Supplementary Table 4). Notably, the apparent low RMSE for PaLM 2 was the result of a constant-output hallucination^14^, where the model produced a limited set of fixed values irrespective of the patient data provided (Supplementary Figure 1). These findings confirm that standalone LLMs are currently not reliable for complex clinical calculations from real-world data.

**Figure 3:**
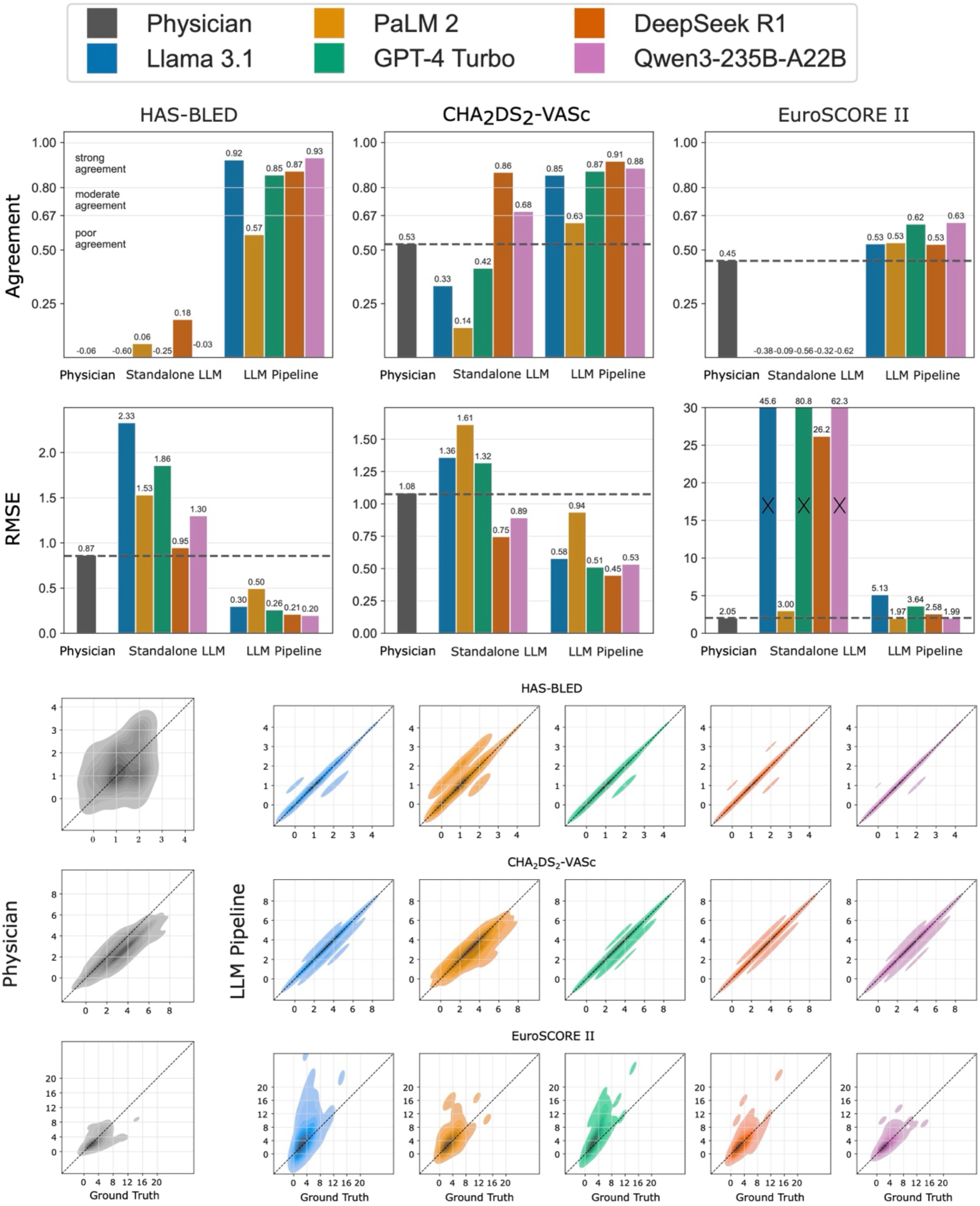
Agreement, error, and calibration. Bar plots represent Krippendorff’s alphas and root mean squared errors (RMSE) for scores derived by treating physicians, standalone LLMs, and LLM pipelines, respectively. Agreement was assessed based on clinically relevant risk score categories^15,16^ i.e., <2 vs ≥2 for CHA_2_DS_2_-VASc^12^, <3 vs ≥3 for HAS-BLED^17^, and <4%, 4–8%, >8% for EuroSCORE II^13^. Calibration is visualized using kernel density estimation plots, with the ground truth score on the x-axis and the predicted score on the y-axis. The diagonal dashed line indicates perfect calibration. Densities closer to this line reflect better calibration between predicted and true scores.

### Knowledge conflicts and algebraic limitations undermine LLM accuracy for complex risk scores

Analysis of per-component accuracies of standalone LLMs (Figure 4) revealed two primary failure modes. First, LLMs often relied on their internal, generalized definitions of medical terms instead of the specific criteria used in the original score publications, a phenomenon known as knowledge conflict^15^ or contextual hallucination^2^. For instance, models frequently assigned a point for ‘Hypertension’ in the HAS-BLED score even when the patient’s blood pressure was well-controlled and did not meet the score’s specific criterion of systolic blood pressure >160 mmHg (Supplementary Table 1). As shown in the radar plots (Figure 4), this led to poor per-component accuracies for several variables.

**Figure 4:**
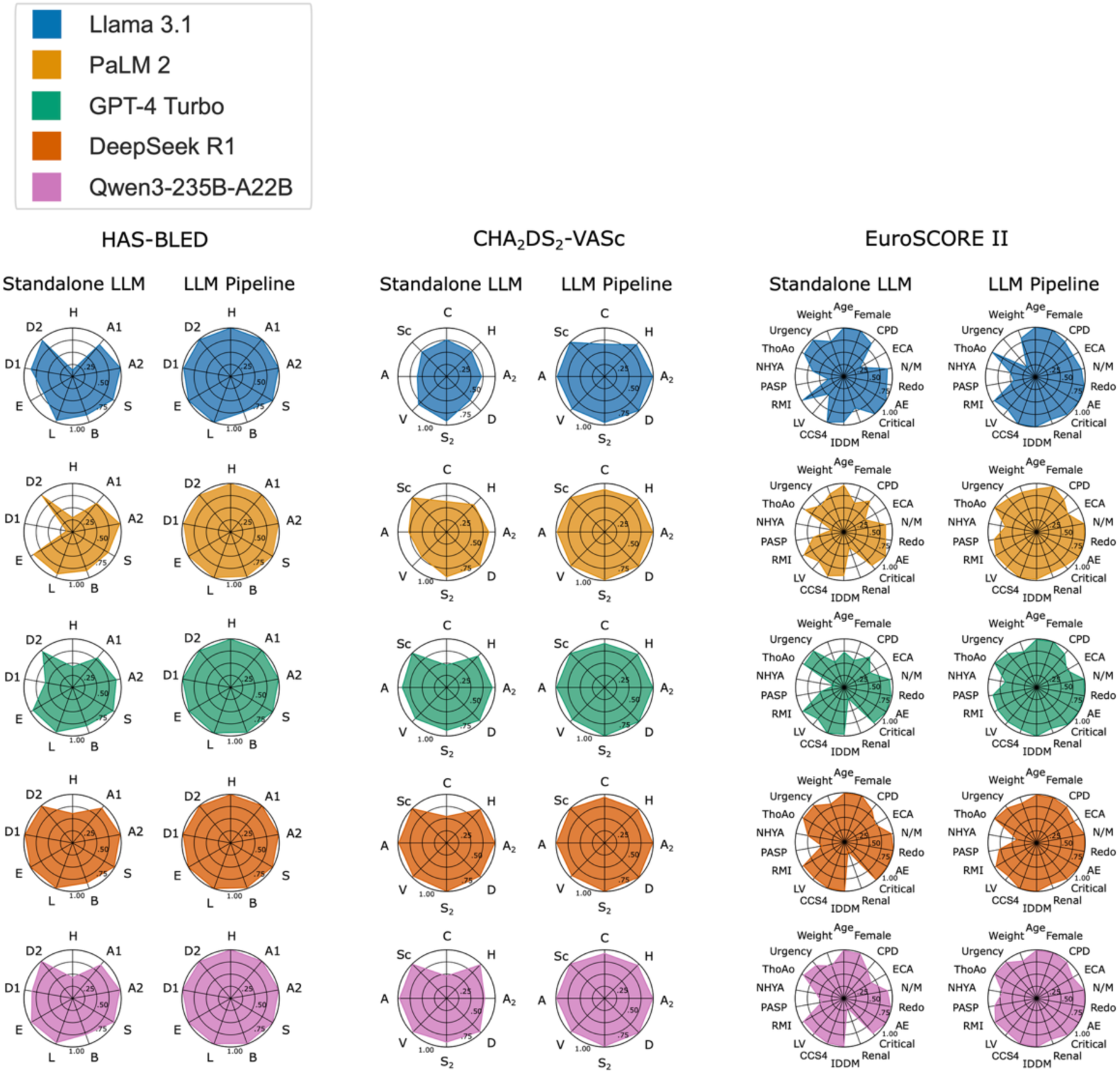
Per-component accuracy. Radar plots represent mean accuracies for individual score components. Mean per-component accuracies across all LLMs are 0.83 (HAS-BLED), 0.85 (CHA_2_DS_2_-VASc), and 0.81 (EuroSCORE II) for the standalone LLM, and 0.99 (HAS-BLED), 0.96 (CHA_2_DS_2_-VASc), and 0.91 (EuroSCORE II) for the LLM pipeline approach. Score component abbreviations are shown in Supplementary Table 3.

Second, the LLMs demonstrated significant algebraic limitations. Models struggled to correctly classify numerical values, such as categorizing a patient’s age or renal function into the appropriate score strata. In an exploratory analysis, PaLM 2 correctly extracted the patient’s age in 98.3% of cases but failed to assign the correct age-related points for the CHA_2_DS_2_-VASc score in 37.4% of those instances. We also frequently observed arithmetic inconsistencies where the sum of component points did not match the final total score reported by the model.

### LLM-based Pipelines Overcome LLM Shortcomings in Algebraic Reasoning

Our pipeline model, which decouples data extraction from score calculation, successfully mitigated these failure modes. Instructing the LLMs to extract only granular data points (e.g., laboratory values, dates, medication data) and delegating all arithmetic and logical operations to a deterministic downstream algorithm substantially improved performance. For most of the LLMs tested, the pipeline approach clearly outperformed both the standalone LLMs and treating physicians across all risk scores and evaluation metrics (Figure 3, Figure 4, Supplementary Table 4). Agreement with the ground truth in clinically relevant categories was markedly higher for the pipeline models compared to scores derived by standalone LLMs and scores documented by treating physicians (mean Krippendorff’s alpha across all scores: 0.79 vs 0.32 vs 0.31). The pipeline models generally achieved lower RMSEs across all scores and demonstrated superior calibration compared to treating physicians. Within the LLM group, models designed for reasoning generally outperformed non-reasoning focused models in both the standalone and pipeline configurations. Notably, treating physicians tended to underestimate patient risk compared to the expert-adjudicated ground truth (Figure 3, Supplementary Table 5).

### Current LLMs are reliable data extractors but may be poor medical data interpreters

Despite the pipeline’s high performance, per-component accuracies remained low for EuroSCORE II variables that require nuanced clinical interpretation, such as NYHA class and urgency. This suggests that while LLMs are reliable at structured data extraction, they are less adept at interpreting ambiguous clinical context. However, these same score components also exhibited the lowest interrater agreement among the expert cardiologists who established the ground truth (Supplementary Table 6). These finding highlights that the challenge in evaluating such variables is rooted in the interpretive nature of their clinical definitions, affecting both humans and artificial intelligence.

### LLM-based pipelines show promise in treatment optimization

In an exploratory analysis, we assessed whether the pipeline could identify discrepancies between guideline-recommended and actual treatment. The DeepSeek-R1-powered pipeline model correctly identified 11 patients in the AF cohort who had a clear indication for anticoagulation (i.e., CHA_2_DS_2_-VASc score ≥2) as well as two patients who might not have had an indication for anticoagulation therapy (i.e., CHA_2_DS_2_-VASc score <2), contrary to the physician’s assessment, respectively (Figure 5). Furthermore, the model identified four patients with documented diabetes mellitus who lacked a prescription for antidiabetic therapy, highlighting the pipeline’s potential to serve as an automated tool for auditing care quality and optimizing treatment.

**Figure 5:**
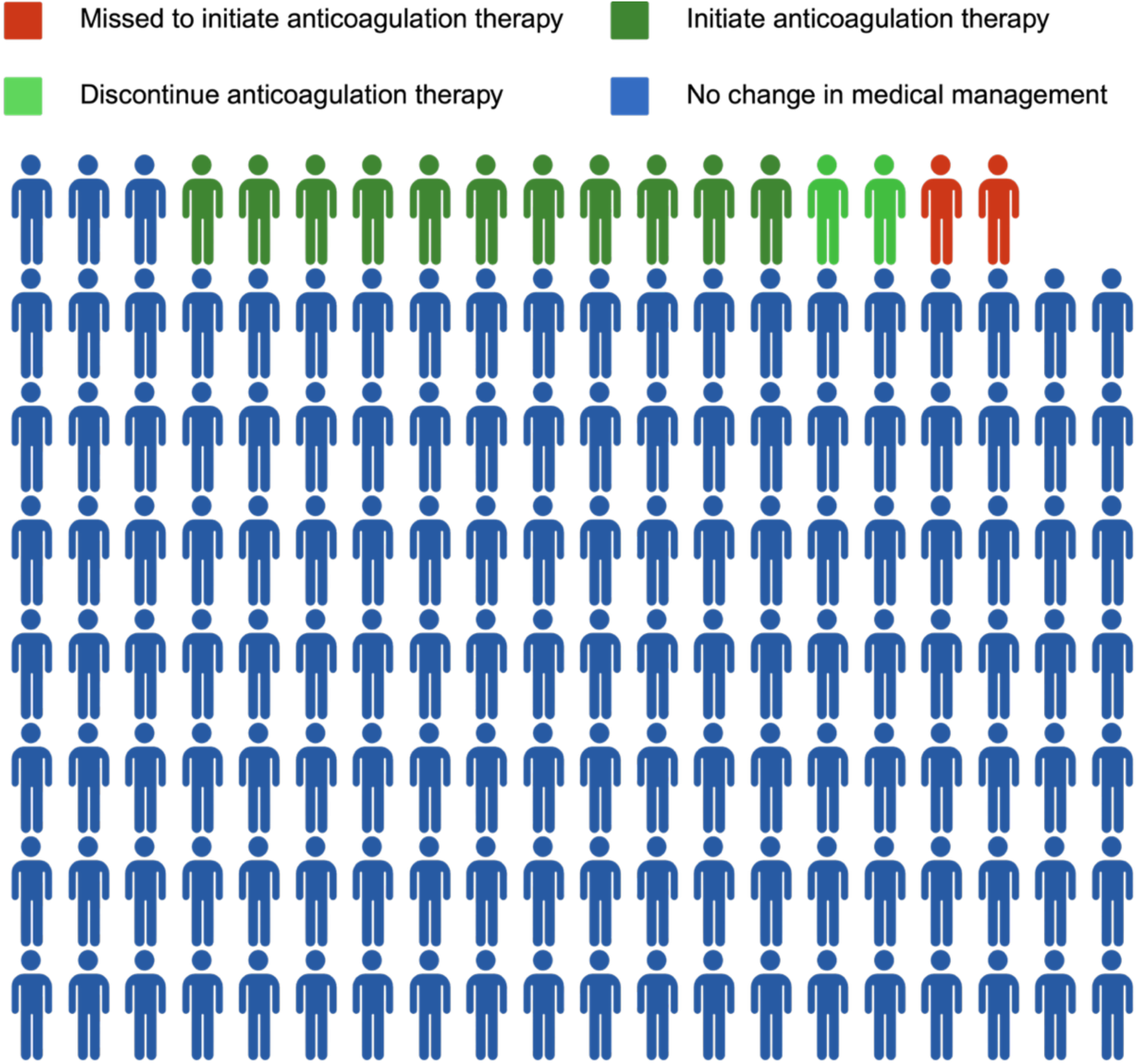
Theoretical clinical impact of re-calculated CHA_2_DS_2_-VASc scores (LLM pipeline using DeepSeek-R1) on anticoagulation management in 142 AF patients: 11 patients (dark green) would have been newly and correctly recommended to initiate anticoagulation therapy, while two patients (light green) would have appropriately had therapy discontinued or at least re-evaluated. Two patients who should have received a recommendation for anticoagulation therapy were missed by both the pipeline model and the treating physicians (red).

## Discussion

In summary, we provide the first evidence that an LLM pipelines can calculate clinical risk scores from unstructured, real-world patient data with an accuracy and calibration superior to that of scores documented by treating physicians. Previous studies have highlighted the potential of LLM-based tools for clinical calculations, but their reliance on synthetic vignettes or simplified case reports limited their clinical applicability. Our work directly addresses the calls from those studies to evaluate performance on authentic clinical documents and against the benchmark of real-world human practice, confirming that a thoughtfully architected pipeline can overcome the known limitations of LLMs in this domain.

A striking finding of our study was the substantial discrepancy between physician-documented scores and the expert-adjudicated ground truth, with treating physicians frequently underestimating patient risk (Supplementary Table 5). While the precise causes are multifactorial, several explanations are plausible. First, clinicians may have an incomplete recall of the specific, nuanced definitions of score components, a known source of miscalculation^16,18^. Second, in fast-paced clinical environments, time constraints may lead to the reuse of outdated scores from previous encounters, a practice that, while pragmatic, contravenes guideline recommendations for regular risk reassessment^12^. Similarly, time constraints in routine clinical practice likely explain the observation that, despite explicit guideline recommendations, the HAS-BLED score was calculated in only 20.1% of patients within the AF cohort. While it could be argued that physicians may have had access to additional clinical information not captured in the written records, this is unlikely to account for the observed discrepancy. Such undocumented risk factors would typically lead to higher calculated scores. In contrast, we observed that the scores documented by treating physicians were, on average, lower than the ground truth scores (Supplementary Table 1, Supplementary Table 2, Supplementary Table 5).

The successful performance of our pipeline models suggests that such systems could be integrated into clinical decision support tools to automate and standardize risk assessment. By decoupling language understanding from mathematical computation, our approach provides a blueprint for leveraging the strengths of LLMs while mitigating their weaknesses. This may remove a key historical barrier in clinical predictive modelling: the reliance on simple, manually calculable risk scores^19^. The ability of LLMs to perform automated, granular data extraction from free-text could unlock the use of more sophisticated and potentially more accurate predictive models that incorporate a far richer set of variables, including narrative-based data that is currently inaccessible to traditional algorithms.

Beyond risk scoring, the pipeline methodology shows promise for broader applications aimed at improving clinical workflow efficiency and patient safety. As our exploratory analysis demonstrated, such a system could be used to audit care and flag potential treatment gaps, such as patients with an indication for anticoagulation or antidiabetic therapy who were not receiving appropriate treatment. This capability could be invaluable for tasks ranging from identifying clinical trial candidates to compiling data for multidisciplinary heart team or tumor board reviews.

### Limitations

Our study has several limitations. The retrospective, single-center design limits generalizability and requires validation in external cohorts. The pipeline depends on expert-crafted prompts, a manual process that can hinder scalability and risks overfitting to institutional documentation. Although emerging frameworks like DSPy^20^ and retrieval-augmented generation (RAG)^21^ may automate prompt engineering and knowledge integration, these remain nascent and often require substantial manual training data and document curation. Finally, although ground truth was established by a rigorous three-expert consensus, it is a proxy, and inherent subjectivity – especially for variables like NYHA class – is a limitation for both human and AI assessment.

## Conclusion

This study provides a validated framework for using an LLM-based pipeline to perform clinical risk scoring from real-world, unstructured data. We show that this approach is not only feasible but can exceed the accuracy of scores documented during routine clinical care. This work lays the foundation for developing robust AI tools and agent-based applications capable of improving the consistency and reliability of clinical risk stratification, ultimately facilitating safer and more guideline-adherent medical practice.

## Code Availability

The analysis pipeline is provided as an MCP server application under github.com/charite-iaim/mcp-risk-server.

## Data availability statement

The (anonymized) data underlying this article will be shared on reasonable request to the corresponding author.

## Supplementary Material

### Supplementary Figures

**Supplementary Figure 1:**
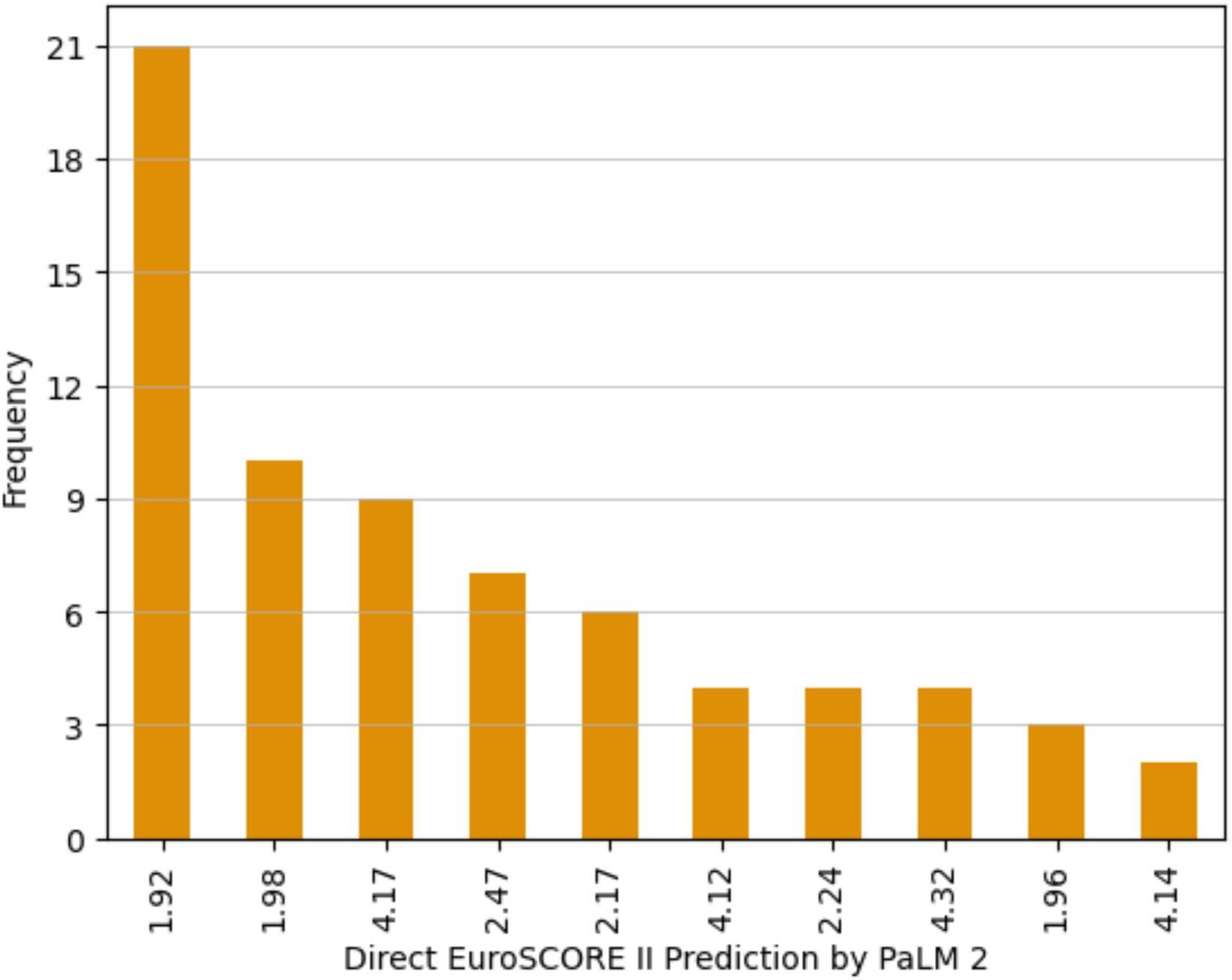
Frequency distribution of EuroSCORE II predictions generated by PaLM 2. The standalone LLM (PaLM 2) produced a limited range of repeated values across heterogeneous patient profiles – suggesting a constant-output hallucination pattern.

### Supplementary Tables

**Supplementary Table 1:** Baseline characteristics of patients with symptomatic atrial fibrillation (n=179) Patient characteristics were reported as counts and percentages for binary variables, as means with 95% confidence intervals (square brackets) for normally distributed continuous variables, and as medians with interquartile ranges (parentheses) for non-normally distributed variables.

| Item | Data availability (%) | Summary statistic |
| --- | --- | --- |
| Age (years) | 100.0 | 69.9 (61.5, 74.5) |
| Documented HAS-BLED score | 20.1 | 1.4 [1.3, 1.5] |
| HAS-BLED score (ground truth) | 100.0 | 1.0 (0.0, 2.0) |
| Uncontrolled hypertension | 100.0 | 13 (7.3%) |
| Renal disease | 100.0 | 0 (0.0%) |
| Liver disease | 100.0 | 0 (0.0%) |
| Stroke history | 100.0 | 17 (9.5%) |
| Bleeding history or predisposition | 100.0 | 25 (14.0%) |
| Labile INR | 100.0 | 0 (0.0%) |
| Elderly (>65 years) | 100.0 | 117 (65.4%) |
| Drugs | 100.0 | 10 (5.6%) |
| Alcohol | 100.0 | 4 (2.2%) |
| Documented CHA<sub>2</sub>DS<sub>2</sub>-VASc score | 79.3 | 2.6 [2.4, 2.8] |
| CHA<sub>2</sub>DS<sub>2</sub>-VASc score (ground truth) | 100.0 | 3.0 (2.0, 4.0) |
| CHF history | 100.0 | 134 (74.9%) |
| Hypertension | 100.0 | 123 (68.7%) |
| Age ≥75 years | 100.0 | 43 (24.0%) |
| Diabetes mellitus | 100.0 | 28 (15.6%) |
| Vascular disease history | 100.0 | 37 (20.7%) |
| Age 65-74 years | 100.0 | 73 (40.8%) |
| Female sex | 100.0 | 67 (37.4%) |
| Systolic blood pressure (mmHg) | 55.9 | 134.5 [131.3, 137.7] |
| LVEF (%) | 77.1 | 55.3 [53.9, 56.7] |
| Hemoglobin (g/dl) | 78.2 | 14.1 [13.9, 14.3] |
| NTproBNP (pg/ml) | 63.1 | 1,264.3 [1044.3, 1484.3] |
| HbA1c (%) | 76.0 | 5.8 [5.7, 5.9] |
| Serum creatinine (mg/dl) | 78.8 | 1.0 [1.0, 1.0] |
| Aspartate transaminase (U/l) | 77.7 | 28.4 [26.7, 30.1] |
| Total bilirubin (mg/dl) | 2.2 | 1.0 [0.9, 1.1] |

**Supplementary Table 2:** Baseline characteristics of patients with severe aortic stenosis (n=76) Patient characteristics were reported as counts and percentages for binary variables, as means with 95% confidence intervals (square brackets) for normally distributed continuous variables, and as medians with interquartile ranges (parentheses) for non-normally distributed variables.

| Item | Data availability (%) | Summary statistic |
| --- | --- | --- |
| Documented EuroSCORE II | 100.0 | 2.5 (1.6, 3.9) |
| EuroSCORE II (ground truth) | 100.0 | 3.1 (1.8, 4.9) |
| Active endocarditis | 100.0 | 0 (0.0%) |
| CCS angina class 4 | 100.0 | 0 (0.0%) |
| Chronic lung disease | 100.0 | 12 (15.8%) |
| Critical preoperative state | 100.0 | 0 (0.0%) |
| Extracardiac arteriopathy | 100.0 | 17 (22.4%) |
| Female sex | 100.0 | 37 (48.7%) |
| Diabetes on insulin | 100.0 | 8 (10.5%) |
| LV function | 100.0 | good: 59 (77.6%)<br>moderate: 14 (18.4%)<br>poor: 3 (3.9%)<br>very poor: 0 (0.0%) |
| Poor mobility | 100.0 | 7 (9.2%) |
| NYHA class | 100.0 | I: 9 (11.8%)<br>II: 22 (28.9%)<br>III: 39 (51.3%)<br>IV: 6 (7.9%) |
| Pulmonary hypertension | 100.0 | 31-55 mmHg: 38 (50.0%)<br>>55mmHg: 5 (6.6%)<br>normal: 33 (43.4%) |
| Recent myocardial infarction | 100.0 | 1 (1.3%) |
| Previous cardiac surgery | 100.0 | 2 (2.6%) |
| Renal impairment | 100.0 | no: 12 (15.8%)<br>cc 50-85: 46 (60.5%)<br>on dialysis: 5 (6.6%)<br>cc <50: 13 (17.1%) |
| eGFR | 90.8 | 64.2 [60.0, 68.4] |
| Surgery on thoracic aorta | 100.0 | 5 (6.6%) |
| Urgency of operation | 100.0 | elective: 74 (97.4%)<br>urgent: 2 (2.6%)<br>salvage: 0 (0.0%)<br>emergency: 0 (0.0%) |
| Weight of operation | 100.0 | isolated CABG 0 (0.0%)<br>single non-CABG: 61 (80.3%)<br>two procedures: 15 (19.7%)<br>three procedures 0 (0.0%) |
| Age (years) | 100.0 | 77.7 [76.1, 79.3] |

**Supplementary Table 3:** Score definitions. Score components, their definitions and the respective points/logistic regression coefficients used in our study are shown for the CHA2DS2-VASc^12^, HAS-BLED^13^ and EuroSCORE II^14^ next to the abbreviations used in Figure 4.

| CHA<sub>2</sub>DS<sub>2</sub>-VASc score |  |  |  |
| --- | --- | --- | --- |
| Score component | Abbreviation | Component definition | Points awarded |
| Congestive heart failure | C | Signs and symptoms of congestive heart failure (i.e., dyspnea at rest or on exertion, fatigue, reduced exercise capacity, dizziness, lightheadedness, peripheral edema) and/or a history of decompensated heart failure and/or a history of tachycardia-induced cardiomyopathy and/or a left-ventricular ejection fraction <40% according to the most recent echocardiography exam. | 1 |
| Hypertension | H | Diagnosis of arterial hypertension and/or current antihypertensive pharmacologic treatment | 1 |
| Age ≥75 y | A<sub>2</sub> | Patient age ≥75 years | 2 |
| Diabetes | D | One or more of the following: Diagnosis of diabetes mellitus, treatment with oral hypoglycemic agents (biguanides, DPP-4 inhibitors, GLP-1 receptor agonists, thiazolidinediones) and/or insulin, HbA1c ≥6.5% | 1 |
| Stroke | S<sub>2</sub> | Prior stroke and/or prior transient ischemic attack and/or a history of arterial thromboembolism | 2 |
| Vascular | V | One or more of the following: Prior myocardial infarction, significant coronary artery disease (defined as a greater than 50% angiographic narrowing of at least one coronary artery and/or a history of percutaneous coronary intervention), peripheral artery disease, complex aortic plaque | 1 |
| Age 65-74 y | A | Patient age between 65 and 74 years | 1 |
| Sex category | Sc | Female sex | 1 |
| HAS-BLED score |  |  |  |
| Score component | Abbreviation | Component definition | Points awarded |
| Hypertension | H | Systolic blood pressure >160 mmHg and/or a diagnosis of uncontrolled hypertension | 1 |
| Abnormal renal function | A1 | One or more of the following: Dialysis, kidney transplant, serum creatinine >200 mmol/L (>2.26 mg/dL) | 1 |
| Abnormal hepatic function | A2 | One or more of the following: Liver cirrhosis, total bilirubin >2 x upper limit of normal, aspartate aminotransferase/alanine aminotransferase/alkaline phosphatase >3 × upper limit normal | 1 |
| Stroke | S | Previous ischemic and/or hemorrhagic stroke | 1 |
| Bleeding | B | Previous major hemorrhage (defined as intracranial bleeding and/or bleeding requiring hospitalization and/or blood transfusion and/or immediate therapy such as endoscopy) and/or anemia (defined as hemoglobin level <13 g/dL if male, and <12 g/dL if female) | 1 |
| Labile INR | L | Time in therapeutic range <60% in patients receiving vitamin K antagonists | 1 |
| Elderly | E | Patient age >65 years | 1 |
| Drugs | D1 | Concomitant use of antiplatelet drugs and/or NSAIDs | 1 |
| Alcohol | D2 | Excessive alcohol consumption defined as >8 alcoholic drinks/week and/or a diagnosis of alcoholism | 1 |

Supplementary Table 3: Score definitions
| EuroSCORE II |  |  |  |
| --- | --- | --- | --- |
| Score component | Abbreviation | Component definition | Regression coefficient |
| NYHA class | NYHA | • I: no symptoms on moderate exertion<br>• II: symptoms on moderate exertion<br>• III: symptoms on light exertion<br>• IV: symptoms at rest | 0<br>0.1070545<br>0.2958<br>0.55979 |
| CCS4 | - | CCS class 4 angina (inability to perform any activity without angina or angina at rest) | 0.2226147 |
| IDDM | - | Insulin-dependent diabetes mellitus | 0.3542749 |
| Age |  | Patient age in years<br>• age $\leq 60$<br>• age $> 60$ | 0.0285181<br>(age - 59)*<br>0.0285181 |
| Female sex | Female | Patient is female | 0.2196434 |
| Extracardiac arteriopathy | ECA | One or more of the following: Claudication, carotid occlusion or $>50\%$ stenosis, prior amputation for arterial disease, previous or planned intervention on the abdominal aorta, limb arteries or carotids. | 0.5360268 |
| Chronic pulmonary disease | CPD | The patient is on bronchodilators or steroids for lung disease | 0.1886564 |
| Poor mobility | N/M | Severe impairment of mobility secondary to musculoskeletal or neurological dysfunction | 0.2407181 |
| Previous cardiac surgery | Redo | One or more previous major cardiac operations requiring opening of the pericardium. | 1.118599 |
| Renal dysfunction | Renal | • CC $>85$ ml/min<br>• CC 50–85 ml/min<br>• on dialysis (regardless of serum creatinine)<br>• CC $<50$ ml/min | 0<br>0.303553<br>0.6421508<br>0.8592256 |
| Active endocarditis | AE | Patients still on antibiotic treatment for endocarditis at the time of surgery. | 0.6194522 |
| Critical preoperative state | Critical | Any one or more of the following occurring preoperatively in the same hospital admission as the operation: ventricular tachycardia or fibrillation or aborted sudden death, cardiac massage, ventilation before arrival in the anaesthetic room, inotropes, intra-aortic balloon counterpulsation or ventricular-assist device before arrival in the anaesthetic room, acute renal failure (anuria or oliguria) | 1.086517 |
| LV function | LV | • good (LVEF $>50\%$)<br>• moderate (LVEF 31–50%)<br>• poor (LVEF 21–30%)<br>• very poor (LVEF $\leq 20\%$) | 0<br>0.3150652<br>0.8084096<br>0.934691 |
| Recent myocardial infarction | RMI | Myocardial infarction within the last 90 days | 0.1528943 |
| PA systolic pressure | PASP | • <31 mmHg<br>• 31-55 mmHg<br>• >55 mmHg | 0<br>0.1788899<br>0.3491475 |
| Urgency | - | • elective: routine admission for operation<br>• urgent: patients not electively admitted for operation but who require surgery on the current admission for medical reasons and cannot be discharged without a definitive procedure<br>• emergency: operation before the beginning of the next working day after decision to operate<br>• salvage: patients requiring cardiopulmonary resuscitation (external cardiac massage) en route to the operating theatre or before induction of anesthesia. This does not include cardiopulmonary resuscitation after induction of anesthesia. | 0<br>0.3174673<br>0.7039121<br>1.362947 |
| Weight of procedure | Weight | • isolated CABG<br>• isolated non-CABG major procedure (e.g. single valve procedure, replacement of ascending aorta, correction of septal defect, etc.)<br>• two major procedures (e.g. CABG + AVR), or CABG + mitral valve repair (MVR), or AVR + replacement of ascending aorta, or CABG + maze procedure, or AVR + MVR, etc.)<br>• three major procedures or more (e.g. AVR + MVR + CABG, or MVR + CABG + tricuspid annuloplasty, etc.), or aortic root replacement when it includes AVR or repair + coronary reimplantation + root and ascending replacement). | 0<br>0.0062118<br>0.5521478<br>0.9724533 |
| Thoracic aorta | ThoAo | Disorder of the thoracic aorta requiring surgery | 0.6527205 |
| Constant |  | - | -5.324537 |

**Supplementary Table 4:** Performance metrics. Krippendorff’s Alpha and root mean squared error (RMSE) for Physician, standalone LLM and LLM pipeline experiments with 95% confidence intervals.

| Score | Experiment | Model | $\alpha$ [CI] | RMSE [CI] |
| --- | --- | --- | --- | --- |
| HAS-BLED | Physician | - | -0.06 [-0.15, 0.00] | 0.87 [0.69, 1.04] |
|  | Standalone LLM | Llama 3.1 | -0.60 [-0.73, -0.46] | 2.33 [2.17, 2.49] |
|  |  | PaLM 2 | 0.06 [-0.12, 0.26] | 1.53 [1.42, 1.64] |
|  |  | GPT-4 Turbo | -0.25 [-0.40, -0.10] | 1.86 [1.66, 2.05] |
|  |  | DeepSeek R1 | 0.18 [-0.03, 0.37] | 0.95 [0.84, 1.06] |
|  |  | Qwen3 | -0.03 [-0.19, 0.14] | 1.30 [1.15, 1.46] |
|  | LLM Pipeline | Llama 3.1 | 0.92 [0.66, 1.00] | 0.30 [0.20, 0.39] |
|  |  | PaLM 2 | 0.57 [0.27, 0.79] | 0.50 [0.43, 0.55] |
|  |  | GPT-4 Turbo | 0.85 [0.56, 1.00] | 0.26 [0.15, 0.35] |
|  |  | DeepSeek R1 | 0.87 [0.60, 1.00] | 0.21 [0.13, 0.28] |
|  |  | Qwen3 | 0.93 [0.71, 1.00] | 0.20 [0.11, 0.26] |
| CHA<sub>2</sub>DS<sub>2</sub>-VASc | Physician | - | 0.53 [0.29, 0.72] | 1.08 [0.96, 1.20] |
|  | Standalone LLM | LLama 3.1 | 0.33 [0.06, 0.58] | 1.36 [1.20, 1.52] |
|  |  | PaLM 2 | 0.14 [-0.06, 0.37] | 1.61 [1.47, 1.75] |
|  |  | GPT-4 Turbo | 0.42 [0.15, 0.65] | 1.32 [1.16, 1.48] |
|  |  | DeepSeek R1 | 0.86 [0.70, 0.97] | 0.75 [0.67, 0.82] |
|  |  | Qwen3 | 0.68 [0.45, 0.85] | 0.89 [0.80, 0.99] |
|  | LLM Pipeline | Llama 3.1 | 0.85 [0.69, 0.97] | 0.58 [0.46, 0.70] |
|  |  | PaLM 2 | 0.63 [0.40, 0.80] | 0.94 [0.81, 1.06] |
|  |  | GPT-4 Turbo | 0.87 [0.73, 0.97] | 0.51 [0.41, 0.61] |
|  |  | DeepSeek R1 | 0.91 [0.79, 1.00] | 0.45 [0.34, 0.54] |
|  |  | Qwen3 | 0.88[0.74, 0.97] | 0.53 [0.40, 0.66] |
| EuroSCORE II | Physician | - | 0.45 [0.18, 0.69] | 2.05 [1.50, 2.56] |
|  | Standalone LLM | Llama 3.1 | -0.38 [-0.57, -0.17] | 45.6 [36.3, 54.1] |
|  |  | PaLM 2 | -0.09 [-0.30, 0.12] | 3.00 [2.26, 3.75] |
|  |  | GPT-4 Turbo | -0.56 [-0.77, -0.34] | 80.8 [75.2, 85.7] |
|  |  | DeepSeek R1 | -0.32 [-0.51, -0.13] | 26.2 [18.6, 33.2] |
|  |  | Qwen3 | -0.62 [-0.77, -0.46] | 62.3 [56.0, 67.9] |
|  | LLM Pipeline | Llama 3.1 | 0.53 [0.31, 0.72] | 5.13 [3.10, 7.06] |
|  |  | PaLM 2 | 0.53 [0.32, 0.71] | 1.97 [1.50, 2.43] |
|  |  | GPT-4 Turbo | 0.62 [0.43, 0.77] | 3.64 [2.71, 4.51] |
|  |  | DeepSeek R1 | 0.53 [0.29, 0.71] | 2.58 [1.61, 3.46] |
|  |  | Qwen3 | 0.63 [0.43, 0.80] | 1.99 [1.41, 2.57] |

**Supplementary Table 5:** Mean error between ground truth and physician-derived scores with 95% confidence intervals. N denotes the number of samples with non-missing scores (i.e., available data) used in the analysis.

| **Score** | **N** | **Mean differences** |
| --- | --- | --- |
| HAS-BLED score | 36 | -0.14 [-0.43, 0.15] |
| CHA<sub>2</sub>DS<sub>2</sub>-VASc score | 142 | 0.64 [0.5, 0.78] |
| EuroSCORE II | 76 | 0.61 [0.16, 1.06] |

**Supplementary Table 6:** Interrater Agreement Metrics Among Clinical Raters. Krippendorff’s alpha is reported for each variable where sufficient variability across categories allowed its calculation. For variables with sparse data—defined as any category represented by a single observation—only raw agreement is presented due to the instability or inapplicability of standard reliability coefficients. Abbreviations as in Supplementary Table 3.

| Variable | Agreement | Raw Agreement |
| --- | --- | --- |
| **HAS-BLED score** | 0.85 | 0.99 |
| Hypertension | 0.88 | 0.98 |
| Abnormal renal function | - | 1.0 |
| Abnormal liver function | - | 0.99 |
| Stroke | 0.64 | 0.96 |
| Bleeding | 0.85 | 0.99 |
| Labile INR | - | 1.0 |
| Elderly (age >65 years) | 0.95 | 0.98 |
| Drugs | 0.88 | 0.99 |
| Alcohol | - | 0.97 |
| **CHA2DS2-VASc score** | 0.97 | 0.99 |
| Congestive heart failure | 0.77 | 0.91 |
| Hypertension | 0.91 | 0.96 |
| Age $\geq 75$ years | 1.0 | 1.0 |
| Diabetes mellitus | 0.95 | 0.99 |
| Stroke | 0.96 | 0.99 |
| Vascular disease | 0.93 | 0.98 |
| Age (65-74 years) | 0.99 | 0.99 |
| Sex category (female) | 1.0 | 1.0 |
| **EuroSCORE II** | 0.94 | 0.97 |
| Active endocarditis | - | 1.0 |
| CCS IV | - | 0.96 |
| Chronic pulmonary disease | 0.72 | 0.92 |
| Critical preoperative state | - | 1.0 |
| Extracardiac arteriopathy | 0.72 | 0.91 |
| Female sex | 0.97 | 0.99 |
| Insulin-dependent diabetes mellitus | 1.0 | 1.0 |
| LV function | 0.90 | 0.95 |
| Poor mobility | - | 0.99 |
| NYHA class | 0.42 | 0.57 |
| PA systolic pressure | 0.81 | 0.88 |
| Recent myocardial infarction | - | 0.97 |
| Previous cardiac surgery | 0.66 | 0.97 |
| Renal dysfunction | 0.68 | 0.96 |
| Thoracic aorta | 0.55 | 0.96 |
| Urgency | 0.25 | 0.88 |
| Weight of procedure | 0.63 | 0.93 |

## References

1. Nateghi R, Aven T. Risk analysis in the age of big data: The promises and pitfalls. Risk Anal. 2021;41(10):1751–1758.

2. Chuang YS, Qiu L, Hsieh CY, Krishna R, Kim Y, Glass J. Lookback lens: Detecting and mitigating contextual hallucinations in large language models using only attention maps. ArXiv Prepr ArXiv240707071. Published online 2024.

3. Khandekar N, Jin Q, Xiong G, et al. Medcalc-bench: Evaluating large language models for medical calculations. Adv Neural Inf Process Syst. 2024;37:84730–84745.

4. Boye J, Moell B. Large language models and mathematical reasoning failures. ArXiv Prepr ArXiv250211574. Published online 2025.

5. Mirzadeh I, Alizadeh K, Shahrokhi H, Tuzel O, Bengio S, Farajtabar M. Gsm-symbolic: Understanding the limitations of mathematical reasoning in large language models. ArXiv Prepr ArXiv241005229. Published online 2024.

6. Goodell AJ, Chu SN, Rouholiman D, Chu LF. Large language model agents can use tools to perform clinical calculations. Npj Digit Med. 2025;8(1):163.

7. Wan N, Jin Q, Chan J, et al. Humans continue to outperform large language models in complex clinical decision-making: a study with medical calculators. ArXiv Prepr ArXiv241105897. Published online 2024.

8. Zhu Y, Wei S, Wang X, Xue K, Zhang X, Zhang S. MeNTi: Bridging medical calculator and LLM agent with nested tool calling. ArXiv Prepr ArXiv241013610. Published online 2024.

9. Lip GY, Nieuwlaat R, Pisters R, Lane DA, Crijns HJ. Refining clinical risk stratification for predicting stroke and thromboembolism in atrial fibrillation using a novel risk factor-based approach: the euro heart survey on atrial fibrillation. Chest. 2010;137(2):263–272.

10. Pisters R, Lane DA, Nieuwlaat R, De Vos CB, Crijns HJGM, Lip GYH. A Novel User-Friendly Score (HAS-BLED) To Assess 1-Year Risk of Major Bleeding in Patients With Atrial Fibrillation. Chest. 2010;138(5):1093–1100. doi:10.1378/chest.10-0134

11. Nashef SAM, Roques F, Sharples LD, et al. EuroSCORE II. Eur J Cardiothorac Surg. 2012;41(4):734-745. doi:10.1093/ejcts/ezs043

12. Hindricks G, Potpara T, Dagres N, et al. 2020 ESC guidelines for the diagnosis and management of atrial fibrillation developed in collaboration with the european association for cardio-thoracic surgery (EACTS) the task force for the diagnosis and management of atrial fibrillation of the european society of cardiology (ESC) developed with the special contribution of the european heart rhythm association (EHRA) of the ESC. Eur Heart J. 2021;42(5):373-498.

13. Vahanian A, Beyersdorf F, Praz F, et al. 2021 ESC/EACTS Guidelines for the management of valvular heart disease: Developed by the Task Force for the management of valvular heart disease of the European Society of Cardiology (ESC) and the European Association for Cardio-Thoracic Surgery (EACTS). Eur Heart J. 2022;43(7):561-632. doi:10.1093/eurheartj/ehab395

14. Greco CM, La Cava L, Tagarelli A. Talking the talk does not entail walking the walk: On the limits of large language models in lexical entailment recognition. ArXiv Prepr ArXiv240614894. Published online 2024.

15. Longpre S, Perisetla K, Chen A, Ramesh N, DuBois C, Singh S. Entity-based knowledge conflicts in question answering. ArXiv Prepr ArXiv210905052. Published online 2021.

16. Zhang J, Lenarczyk R, Marin F, et al. The interpretation of CHA2DS2-vasc score components in clinical practice: a joint survey by the european heart rhythm association (EHRA) scientific initiatives committee, the EHRA young electrophysiologists, the association of cardiovascular nursing and allied professionals, and the european society of cardiology council on stroke. EP Eur. 2020;23(2):314–322. doi:10.1093/europace/euaa358

17. Pisters R, Lane DA, Nieuwlaat R, De Vos CB, Crijns HJ, Lip GY. A novel user-friendly score (HAS-BLED) to assess 1-year risk of major bleeding in patients with atrial fibrillation: the Euro Heart Survey. Chest. 2010;138(5):1093–1100.

18. Zhang C, Shen L, Pan MM, Zheng YL, Gu ZC, Lin HW. Perceptions and knowledge gaps on CHA2DS2-VASc score components: a joint survey of Chinese clinicians and clinical pharmacists. Postgrad Med. 2022;134(1):64–77. doi:10.1080/00325481.2021.1996815

19. Sachs MC, Sjölander A, Gabriel EE. Aim for clinical utility, not just predictive accuracy. Epidemiol Camb Mass. 2020;31(3):359–364.

20. Khattab O, Singhvi A, Maheshwari P, et al. Dspy: Compiling declarative language model calls into self-improving pipelines. ArXiv Prepr ArXiv231003714. Published online 2023.

21. Lewis P, Perez E, Piktus A, et al. Retrieval-augmented generation for knowledge-intensive nlp tasks. Adv Neural Inf Process Syst. 2020;33:9459–9474.

